# Optical genome mapping identifies clinically relevant somatic structural variation in epilepsy-affected brain tissue

**DOI:** 10.64898/2026.04.18.26350985

**Authors:** Anthony R. Miller, James J. Anderson, Maria Elena Hernandez Gonzalez, Lakshmi Prakruthi Rao Venkata, Eileen Stonerock, Lauren Mashburn-Warren, Allison Daley, Jeffrey Leonard, Jonathan Pindrik, Ammar Shaikhouni, Daniel R. Boué, Diana L. Thomas, Christopher R. Pierson, Adam P. Ostendorf, Elaine R. Mardis, Daniel C. Koboldt, Katherine E. Miller, Tracy A. Bedrosian

## Abstract

Somatic variants are a prominent cause of epilepsy-associated cortical malformations, but about half of patients undergoing genetic testing have no finding due partly to limitations in variant detection. Most studies have focused on single-nucleotide variants or small indels that are accessible to short-read sequencing technologies, but somatic structural variants are also emerging as important contributors despite their unique detection challenges. Optical genome mapping (OGM) is a promising methodology for the detection of structural variants, but requires high quality, high molecular weight DNA from clinical specimens. Here we successfully optimize a protocol for OGM of surgically-resected patient brain tissue which yields ∼450x effective coverage – suitable for detecting somatic variants at low allele fractions. We apply this approach to brain specimens from four patients with epilepsy. OGM identifies large and complex mosaic structural variants ranging from 7-40% variant allele fraction, most of which are not captured by short-read exome sequencing of the same specimen. In one patient with a known germline *DEPDC5* variant, OGM reveals a somatic variant *–* a 13.2kb deletion in *DEPDC5* at approximately 20% VAF – consistent with the established two-hit model in DEPDC5-associated lesional epilepsies. By resolving the breakpoints in PacBio HiFi sequencing data, we identify a mechanism for this somatic deletion, mediated by recombination of two Alu elements flanking the region. Our findings demonstrate that OGM is a robust and complementary tool for detecting somatic structural variation in human brain tissue, with potential to improve diagnostic yield and refine genotype–phenotype correlations in neurological disorders.

## Introduction

Somatic mosaicism is a fundamental feature of the human brain, where individual cells differ from one another at the genomic level. This genomic diversity first arises early in development from somatic variants that occur after fertilization and then continue to accumulate during brain development (Rohrback et al. 2018). As cells divide and differentiate, replication errors, DNA damage, and mobile element activity have the potential to introduce a wide range of genomic alterations including single-nucleotide variants, small insertions and deletions, copy number changes, mobile element insertions, and structural variants (Sran et al. 2024). These mutations are inherited by daughter cells, creating clonal lineages with distinct genetic signatures. The resulting mosaicism contributes to the cellular heterogeneity of the human brain with potential roles in shaping neural function and disease susceptibility. Brain somatic variants have been directly tied to several neurological conditions, including developmental brain disorders (Corrigan et al. 2025).

Malformations of cortical development and other developmental lesions are a major cause of pediatric drug-resistant epilepsies referred to surgical intervention (Blumcke et al. 2017). Through next-generation sequencing of resected brain tissue, somatic variants have been identified as an important cause of epilepsy-associated cortical malformations; however, most efforts to detect brain somatic variants in epilepsy thus far have relied on short-read exome sequencing which is limited in its ability to detect large or complex structural variants (SVs). Perhaps due to this focus on protein-coding single nucleotide variants (SNVs) and small indels, only about half of patients undergoing genetic testing of surgically resected brain tissue have a causal somatic variant identified (Bedrosian et al. 2022; Lai et al. 2022; Chung et al. 2023; Lopez-Rivera et al. 2023). This is particularly relevant because mTOR-pathway epilepsies with germline variants in GATOR1 (*DEPDC5/NPRL2/NPRL3*) or *TSC1/2* often follow a two-hit model in which a somatic second hit arises in the lesional cortex (Ribierre et al. 2018). Recently, one study applied deep genome sequencing to frozen brain tissue from 21 unsolved cases of focal epilepsy. Approximately one-fifth of those cases were positive for a disease-causing germline SV that had been missed by other approaches (Deng et al. 2024). SVs in the somatic setting, however, remain mostly unexplored, but emerging data supports their role in focal seizures. In recent studies, we and others identified mosaic chr1q copy number gains as a recurrent cause of focal epilepsies (Kobow et al. 2020; Lopez-Rivera et al. 2023; Miller et al. 2023; Leng et al. 2024). Other somatic copy number variants have been reported in individual cases of epilepsies with cortical malformations (Vasudevaraja et al. 2021; Lopez-Rivera et al. 2023). Collectively, the recent literature suggests that SVs underlie a significant proportion of focal epilepsy, but these variants are rarely interrogated in surgical tissue.

Optical genome mapping (OGM) recently emerged as a highly sensitive, amplification-free and digestion-free genomic technology capable of detecting structural variants across the entire genome, including those in complex or repetitive regions. OGM uses an enzymatic reaction to fluorescently label the genome at a recurring sequence motif. The labeled DNA molecules are linearized and visualized to identify changes in patterns that may reflect sequence variants. This technique offers single-haplotype resolution, enabling identification of large structural alterations and somatic non-coding variants that are often missed by short-read sequencing technologies. Therefore, OGM provides a unique opportunity to profile somatic structural variation in the human brain. To date, the main clinical applications of this technology are somatic profiling of hematological malignancies using patient leukocytes, solid tumors, and constitutional genomic testing (Neveling et al. 2021; Sahajpal et al. 2021; Pang et al. 2023; Sahajpal et al. 2023; Levy et al. 2025). However, the use of OGM in non-tumor brain tissue remains largely underexplored. Here we optimized a protocol for OGM of brain specimens obtained from epilepsy surgery, which produced the first optical genome maps of non-tumor human brain tissue and facilitated the identification of a pathogenic somatic structural variant in a patient with focal epilepsy.

## Results

### Optimized protocol for OGM of patient brain samples

OGM is commonly used for structural variant detection in the germline or in tumor tissue (Levy et al. 2025); however, no studies have applied this approach to examine non-malignant human brain samples. We thus began by optimizing an OGM protocol for human brain tissue. Bionano offers two extraction protocols for solid tissue: ‘SP Tissue and Tumor’ and ‘SP Brain Tissue’. Because DNA length and quality are critical for OGM, we first compared the yields of both protocols on an ‘ideal’ sample type: high-quality, snap frozen mouse brain tissue. We divided the tissue into two equal 15 mg portions (the recommended input for the Brain Tissue protocol) for parallel processing with each method. Both protocols yielded high-molecular weight DNA of comparable quality (N50 >150kb, ∼213–223kb). However, the Brain Tissue protocol produced a higher overall yield (5,173 ng vs. 3,170 ng) with similar effective coverage (471x vs. 502x) and map rates (82.9% vs. 86.5%). Given these results, we proceeded with the Bionano Brain Tissue DNA isolation protocol for our patient brain specimens **(Fig. 1).**

**Fig. 1:**
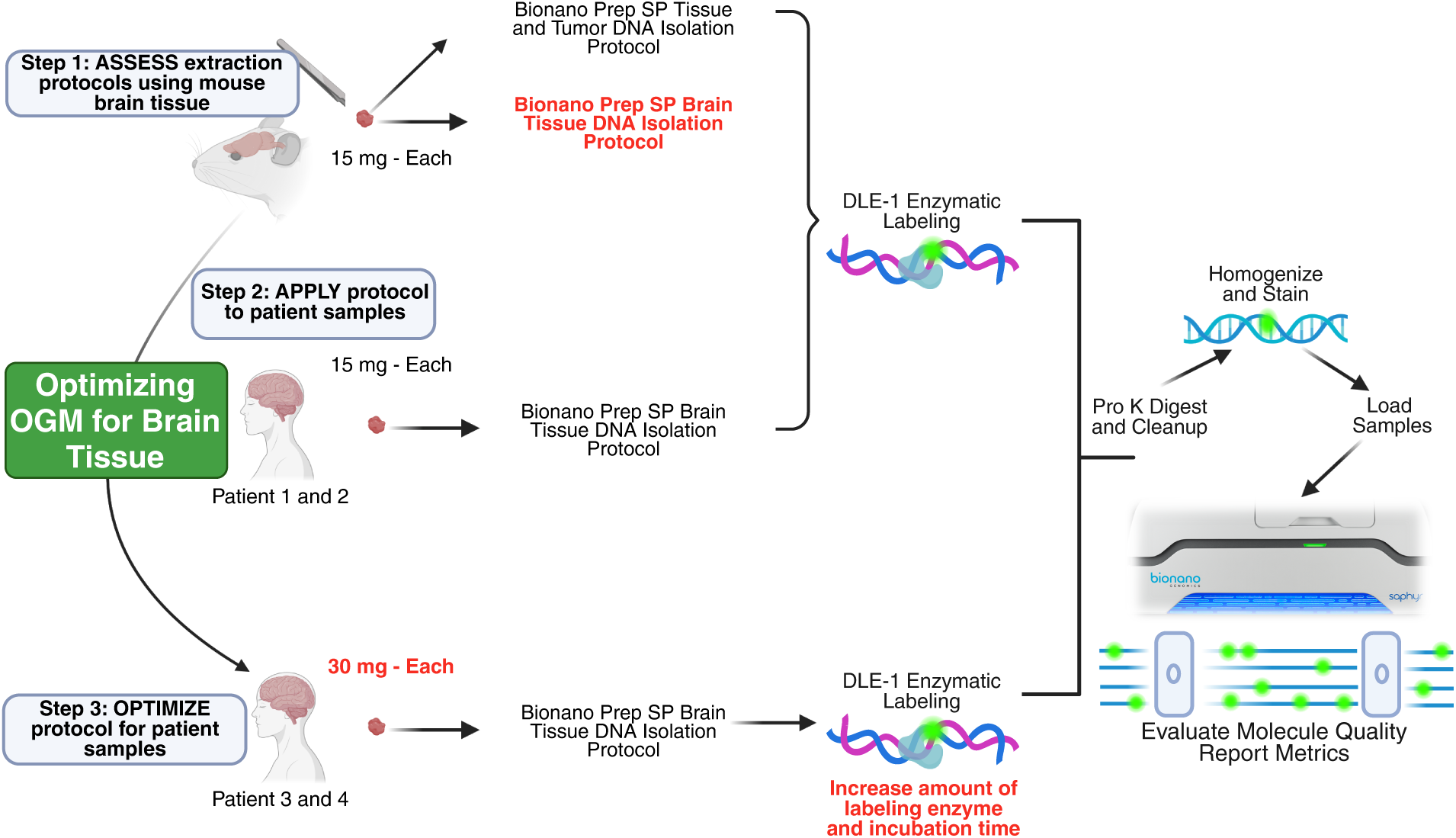
Optimized protocol for optical genome mapping of brain tissue. Workflow demonstrating the preparation of tissues and nucleic acids. Variables tested are highlighted in red.

### Cohort and prior testing

We analyzed frozen surgical specimens from four patients with drug-resistant lesional focal epilepsy **(Table 1).** Three patients carried pathogenic/likely pathogenic germline variants in *NPRL3*, *DEPDC5*, or *TSC1*. All four had prior short-read exome sequencing of resected brain (and blood) that did not reveal any disease-associated somatic variants.

**Table 1:** Patient / Sample Data. Patient brain samples obtained from epilepsy surgery. Histopathological diagnosis was determined by a board-certified neuropathologist following ILAE guidelines. Germline genetic findings were identified through exome sequencing and evaluated according to ACMG criteria with follow-up parental testing when possible. Exome findings from patients 1 and 2 are previously published in (Bedrosian et al. 2022).

| Patient | Sex | Age Range (years) | Brain region | Histopathological diagnosis | Germline genetic findings | Zygosity | Origin |
| --- | --- | --- | --- | --- | --- | --- | --- |
| 1 | M | 11-15 | Right frontal cortex | FCD 1c | *NPRL3*<br>c.995_997del:p.Asn332Thrfs*80 | Heterozygous | Unknown |
| 2 | M | 0-5 | Right frontal lobe | FCD 1c | *DEPDC5* c.346C > T:p.Arg116* | Heterozygous | Paternally inherited |
| 3 | M | 0-5 | Right frontal lobe | Cortical tubers | *TSC1* c.1498C>T:p.Arg500* | Heterozygous | De novo |
| 4 | F | 6-10 | Left hippocampus | Hippocampal sclerosis | None | N/A | N/A |

We first applied OGM to surgically resected frozen brain specimens from two patients (Patients 1 and 2), initially starting with 15 mg of tissue input into the Brain Tissue DNA isolation protocol. Relative to mouse tissue, extraction yields from patient specimens were lower (1,911-2,247 ng) with lower effective coverage (181-393x) and map rate (68.4-79.1%). This is likely related to tissue quality and handling limitations inherent to surgical samples. Therefore, we adjusted our approach by doubling the starting material to 30 mg tissue, increasing the amount of DLE-1 labeling enzyme from 3.0 to 3.5 ul, and extending the labeling time from 1 to 2 hours to address the poor yield and map rate. In the two subsequent 30 mg specimens, total DNA yield increased to 4,231-4,365 ng, effective coverage to 421-458x, and map rates to 82.8-91.0%; these improvements reflected recovery of longer DNA, which closely tracked with obtained coverage, while normalized extraction efficiency remained similar (15 mg: 127-150 ng/mg; 30 mg: 141-146 ng/mg) **(Table 2).** Overall, the protocol modifications resulted in optimized data from patient brain specimens consistent with results obtained from high-quality mouse brain tissue.

**Table 2:** Data output and quality metrics for patient brain samples. Frozen, non-tumor brain specimens from four epilepsy surgical resections were processed on the Bionano Saphyr using the Brain Tissue DNA isolation protocol and DLE-1 labeling. Columns report: **Tissue storage duration (years); Extraction yield (ng/mg)** (Qubit DNA mass normalized to tissue input); **% >60 kb (gDNA TapeStation)** (fraction of fragments >60 kb by Agilent ScreenTape); **NG50 (>150 kb)** (long-molecule N50 computed on molecules ≥150 kb); **Total DNA >150 kb & min sites >9 (Gb)** (gigabases of molecules ≥150 kb carrying ≥9 DLE-1 labels—i.e., usable long molecules); **Avg label density (per 100 kb)** (mean DLE-1 labels per 100 kb; typical target ∼14–17/100 kb); **Effective coverage (x)** (aligned long-molecule bases divided by haploid genome size); **Map rate (%)** (percentage of molecules that align to the reference); and **Positive/Negative label variance (%)** (dispersion of over-/under-labeling relative to expected motif density; lower indicates more uniform labeling). Patients 1–2 used 15 mg tissue input; Patients 3–4 used 30 mg.

| Patient | Tissue storage duration (years) | Extraction yield (ng/mg) | % >60kb (gDNA tapestation) | NG50 (>150kb) | Total DNA >150 kb & min sites >9 (Gb) | Avg Label Density | Effective Coverage | Map Rate | Positive Label Variance | Negative Label Variance |
| --- | --- | --- | --- | --- | --- | --- | --- | --- | --- | --- |
| 1 | 4.5 | 149.8 | 80.0 | 199.1 | 815.8 | 17 / 100 kb | 181x | 68.40% | 3.00% | 9.80% |
| 2 | 7.6 | 127.4 | 94.6 | 198.8 | 1535.8 | 15.4 / 100kb | 393x | 79.10% | 4.20% | 9.20% |
| 3 | 2.8 | 145.5 | 97.8 | 237.5 | 1570.8 | 15.8 / 100 kb | 421x | 82.80% | 5.70% | 7.80% |
| 4 | 2.5 | 141.0 | 97.9 | 257.6 | 1553.4 | 15.4 / 100kb | 458x | 91.00% | 3.80% | 8.20% |

### OGM detects somatic structural variants in human brain

Recognizing that somatic mosaicism is a key feature of all human brains, we initially sought to explore the landscape of clonal somatic structural variation identifiable by OGM regardless of pathogenicity. We analyzed the four patient brain datasets **(Fig. 2A)** using the Bionano Rare Variant Analysis pipeline, initially filtering for variant allele fractions <40% for autosomal chromosomes and <90% for male X chromosomes to enrich for probable somatic variants. In addition, we filtered for variants present in <10% of the Bionano control database, a collection of variants from human blood genomes of diverse ethnic composition without reported disease phenotypes, reasoning that true somatic variants would be relatively rare on a population level. This analysis identified 34 variants overall, or 6-12 variants per individual, representing mostly deletions and insertions as well as four duplications and one inversion **(Supplemental Table S1, Fig. 2B)**. Of these 34 variants, 10 were present in <1% of the control database and 4 variants were absent from the control database entirely. Variant allele fractions ranged from 7-40% with a mean of 30.4% **(Fig. 2C).** Over 70% of variants were less than 30 kb in length **(Fig. 2D).** Variants demonstrated no significant enrichment on any individual chromosome, suggesting uniform distribution across the genome (Binomial test with Bonferroni’s correction for multiple testing, adj. p>0.05, **Fig. 2E).** Most variants spanned both intragenic and intergenic regions, with many overlapping regions containing regulatory elements such as promoters and enhancers **(Fig. 2F).** Notably, several variants exhibited a composition of short or long interspersed nuclear elements (SINEs and LINEs) more than two-fold higher than the genomic average **(Fig. 2F).** To evaluate the statistical significance of these observations, we performed permutation testing (n_perm_= 10,000), which confirmed significant enrichment for three of four likely SINE-enriched variants (p < 0.05). To confirm structural variants identified by OGM, we performed PacBio HiFi sequencing on the same DNA extracted for OGM and reviewed the long-read data visually using Integrated Genomics Viewer. This approach allowed us to orthogonally confirm 22/34 structural variants (65%). The type and size of structural variant influenced the validation rate: we visualized 81% of deletions, 54% of insertions, and 50% of duplications (see Supp. Table 1). Limitations of the PacBio data, including lower coverage relative to OGM, may explain why some variants were not able to be visualized. To assess the possible functional importance of all variants, including both non-coding and coding variants, we evaluated their overlapping and nearest non-overlapping protein-coding genes for inclusion in various disease gene lists. Several variants affected genes that are associated with epilepsy or neurodevelopmental disorders **(Fig. 2G)** and/or were present in regions predicted to be pathogenic by AnnotSV in the context of constitutional disorders **(Fig. 2H).**

**Fig. 2:**
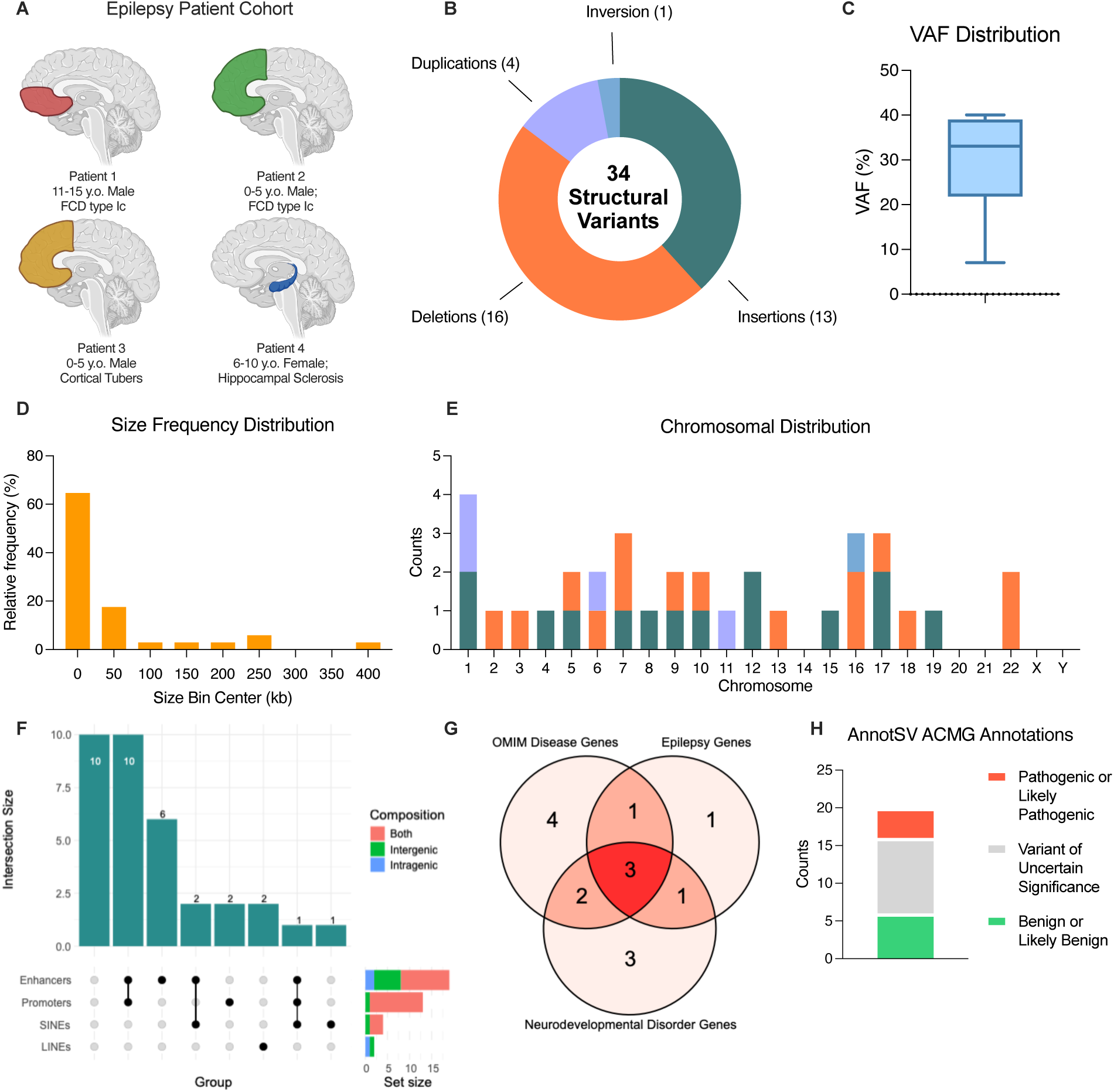
Rates and spectra of brain somatic variants identified by OGM. **(A)** Brain tissues from four epilepsy surgery patients, all affected by developmental brain lesions, were used for OGM. **(B)** Following manual filtering for data quality, OGM identified a set of 34 SVs across the four patient samples, containing mostly deletions and insertions, but also four duplications and one inversion. **(C)** The distribution of variant allele frequencies ranged from 7-40%, with a median of 33%. **(D)** Approximately 65% of the variants in our set were between 0-25 kb in size, though the remaining variants ranged widely, even containing a variant as large as 401 kb. **(E)** The SVs were dispersed throughout the genome with no obvious predisposition for a chromosomal location. **(F)** Annotation of genomic features within SV reference regions revealed overlap with regulatory clusters (≥ 5 enhancers or ≥ 1 promoter), and likely enriched for repetitive elements (≥ 2-fold genomic mean for SINEs and LINEs). Notably, over 70% of our variants occurred in regions likely enriched for at least one of these features. **(G)** Many of the protein coding, overlap and nearest non-overlap genes were also found in gene sets associated with disease, including neurodevelopmental disorders and epilepsy. **(H)** Annotation results from AnnotSV predict that 5% of the variants with sufficient annotation evidence in our set are pathogenic or likely pathogenic in the setting of constitutional disorders.

### OGM identifies a pathogenic somatic second hit in DEPDC5

In response to these findings associating several candidate events with neurological disease, we then applied a strict examination of all structural variants to classify disease relevance. Of all somatic variants identified by OGM and PacBio’s structural variant calling tool, only one event (an intragenic *DEPDC5* deletion in Patient 2) was considered disease relevant as it was absent from the Bionano control database, identified as an epilepsy-associated gene, and was scored Pathogenic by AnnotSV. No other relevant variants were found by OGM or PacBio in the other three patients. Patient 2 carries a heterozygous germline loss-of-function variant in *DEPDC5* previously identified by exome sequencing (p.Arg116*) and Pathogenic by ACMG criteria (Richards et al. 2015). Sanger sequencing validated the germline variant, and additional sequencing of parental samples showed that it was inherited from the unaffected father **(Fig. 3A).** The somatic structural variant identified by OGM was a large deletion within *DEPDC5* at 20% variant allele fraction **(Fig. 3B).** This variant deletes the first several exons of *DEPDC5* and is therefore expected to abolish protein function, consistent with the two-hit model for GATOR1-related focal cortical dysplasia.

**Fig. 3:**
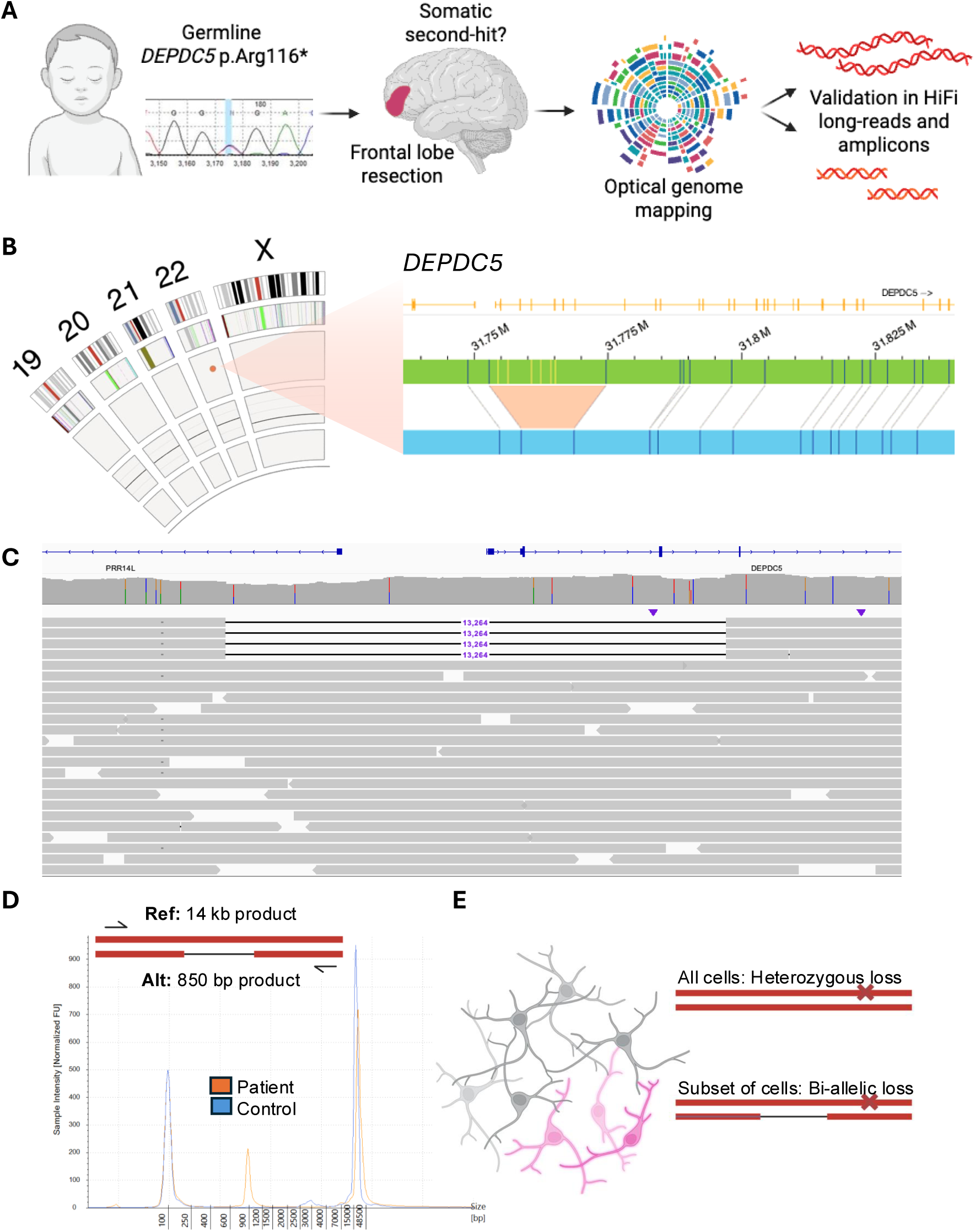
OGM detects somatic *DEPDC5* second hit missed by exome sequencing. **(A)** Exome sequencing initially identified a paternally inherited germline *DEPDC5* truncating variant in an individual treated for focal epilepsy and cortical dysplasia. No relevant somatic variants were identified in this patient by exome sequencing of affected brain tissue. Optical genome mapping of DNA from surgically resected brain tissue was used to search for a potential somatic second hit which would be expected based on clinical presentation. **(B)** BioNano Access view of optical genome mapping results, which identified a somatic mosaic deletion affecting *DEPDC5.* **(C)** IGV view of PacBio HiFi sequencing reads, which validated the 13.2 kb deletion and resolved the precise breakpoints. **(D)** Long-range PCR followed by Sanger sequencing further confirmed the variant and expected breakpoints. **(E)** The somatic deletion was observed on the opposite allele from the patient’s germline variant in HiFi reads, suggesting that a subset of cells carry bi-allelic loss of *DEPDC5*.

Visual review of the PacBio long-read data from Patient 2’s brain tissue revealed a somatic deletion of 13.2 kb (chr22: 31,747,053 - 31,760,316) that was represented by 4 reads and overlapped with the variant called by OGM **(Fig. 3C).** Coverage in this region ranged from 34x-42x. Notably, this variant was not called by PacBio software, likely due to the relatively few supporting reads, highlighting an advantage of OGM for variant discovery in the somatic context. Furthermore, both breakpoints were present in introns, making it challenging to detect using short-read exome sequencing approaches. On the other hand, the variant breakpoints identified by OGM were different from what was observed in PacBio data, demonstrating an advantage of PacBio sequencing for single-base resolution whereas OGM resolution is limited by label density and coverage.

To rule out any potential artifacts of the library preparation process, we performed an additional validation by amplifying the deleted region using long-range PCR with primers flanking the breakpoints. Reference DNA from an unaffected control individual showed the predicted 14kb band, whereas patient DNA showed, as predicted, two products: the 14kb band and an 850bp band representing the deletion **(Fig. 3D).** Sanger sequencing of the 850bp product further confirmed the deletion and its breakpoints.

Notably, three of the four HiFi reads supporting the deletion spanned all the way to the patient’s germline heterozygous variant located in exon 6. None of these three reads contained the germline variant, suggesting that the somatic structural variant is *in trans* with the germline variant affecting the other allele, and thus resulting in bi-allelic inactivation of *DEPDC5* in affected cells **(Supplemental Fig. S1, Fig. 3E).** Taken together, these results verify the somatic second hit in *DEPDC5* identified by OGM and provide a genetic explanation for the patient’s focal seizures and cortical dysplasia.

### Recombination of Alu elements mediates the DEPDC5 somatic variant

To gain insight into the mechanism triggering the *DEPDC5* somatic deletion, we investigated its genomic context for potential clues. Interestingly, two AluY SINE elements are reported in the reference genome at the locations of the breakpoints **(Fig. 4A).** Alu elements are one of the most abundant retrotransposon families in the human genome and have the capacity to generate genomic instability through intrachromosomal and interchromosomal recombination events owing to their high sequence homology (Ade et al. 2013). The presence of these elements at the verified breakpoints of the deletion supports the hypothesis that the *DEPDC5* somatic deletion occurred through Alu-mediated recombination **(Fig. 4B).** The event likely occurred in a neural progenitor cell undergoing cell division during prenatal development, thus leading to a 10-20% variant allele fraction in affected brain regions.

**Fig. 4:**
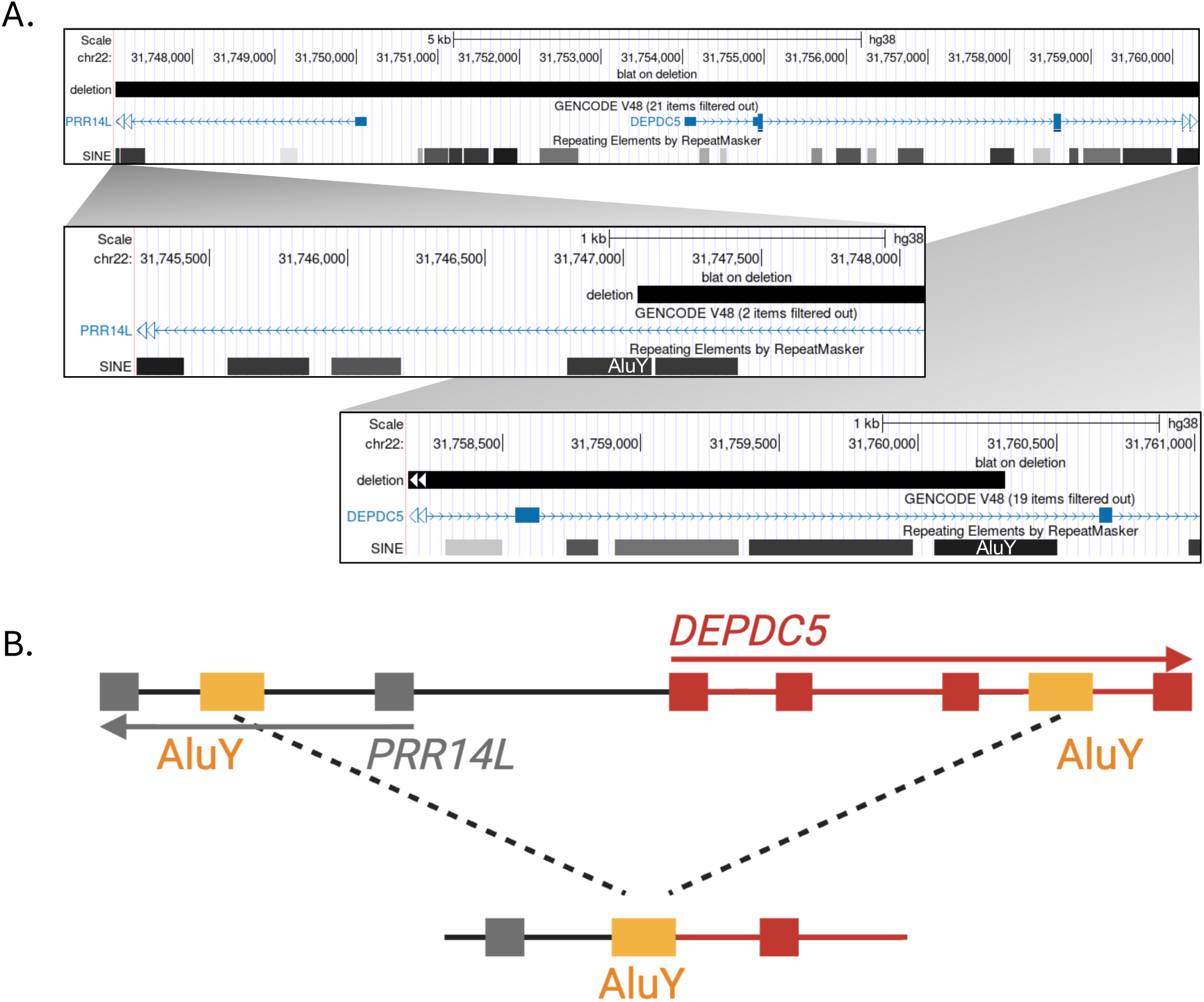
*DEPDC5* variant likely occurred by somatic recombination of Alu elements. **(A)** UCSC genome browser view showing the deleted region with AluY SINE elements overlapping the breakpoints. **(B)** Proposed schematic showing recombination of AluY elements resulting in the observed deletion.

## Discussion

OGM has emerged as a useful technology for identifying structural variation that is difficult or costly to detect with other sequencing-based methodologies. Most applications to date have focused on blood, bone marrow aspirates, and solid tumor specimens whereas none, to our knowledge, applied OGM to non-tumor human brain tissue (Levy et al. 2025). Here we provide evidence for simple modifications to existing protocols that result in high quality OGM data from patient brain specimens. One limitation is the scarcity of tissue available to test all protocol modifications on the same samples in parallel, which is a common limitation in working with patient samples. Tissues from patients 3 and 4 had shorter storage durations, label densities ∼15-16/100 kb, and originated from lower-myelin regions (hippocampus or cortical tuber); these factors may have contributed to their improved metrics, though larger cohorts are needed to draw firm conclusions regarding surgical tissues and their performance. Nevertheless, our results showed consistently improved metrics with increased tissue input, labeling enzyme, and labeling time.

By leveraging these datasets, we generated the first optical genome maps of human brain tissue, which revealed large, clonal somatic structural variants that would be otherwise difficult to detect with alternative sequencing-based approaches. Somatic structural variants are prevalent in human brain tissue with at least 10% of cortical neurons containing large scale copy number variants according to single-nuclei genome sequencing studies (Rehen et al. 2001; McConnell et al. 2013; Sekar et al. 2020). However, these types of studies are costly and limited in terms of the numbers of nuclei sequenced, while requiring genome amplification which can introduce artifacts. Our results using an amplification-free approach show that large clonal mosaic structural variants are present in human brain tissue and confirm that large deletions are more prevalent than large duplications, as previously published (McConnell et al. 2013). Our data provide orthogonal evidence for the presence of mosaic structural variation affecting many cells in human brain tissue.

Identification of true mosaic structural variants is subject to caveats around filtering criteria and the lack of OGM data on a matched germline sample. We selected a <10% allele frequency threshold in the Bionano control database to balance sensitivity for detecting somatic events with exclusion of common polymorphic variants. Because OGM breakpoints are approximate rather than exact, a 10% allele frequency in the control database does not necessarily indicate that an identical structural variant is present in 10% of individuals. Reported frequency can reflect recurrent variation anywhere within that broader genomic interval rather than the same precise event. Many bona fide somatic structural variants, particularly those arising in mosaic contexts, occur in regions that are structurally variable in the population and would be excluded by a 1% cutoff. Therefore, we used a less stringent 10% filter in this analysis to help retain potentially relevant somatic events that would be excluded under germline pathogenicity filters designed for rare constitutional disease variants, but conversely, this could result in unintended inclusion of some rare germline polymorphisms. In addition, we used a VAF filter of <40% to enrich for mosaic variants; however, VAF alone is not sufficient to distinguish germline from somatic structural variants, particularly in the context of OGM, where quantitative estimates are influenced by molecule sampling and alignment biases. Nevertheless, somatic mosaic variants arising early in development may be present at allele fractions approaching 50%. These caveats must be considered in the absence of a matched germline comparator sample for each individual.

On the other hand, for detecting disease-relevant variants, we used a filter of <1% allele frequency in the population, which aligns with field standards for detecting pathogenic variation. Using this cutoff, OGM allowed for the identification of a pathogenic somatic variant previously missed by other approaches. It is possible that the *DEPDC5* variant could have been initially identified by PacBio sequencing alone; however, the variant allele fraction was near the lower limit of sensitivity for this technology and was not called using default analysis settings. There are strengths and limitations to both approaches. OGM provides greater sensitivity for mosaic variants and superior ability to resolve large or complex structural variants at lower cost. On the other hand, PacBio provides better resolution of breakpoint sequences, as observed in our confirmatory studies, thus making the two technologies complementary for somatic structural variant detection.

In conclusion, our findings demonstrate that OGM is a robust and complementary tool for detecting mosaic structural variation in human brain tissue. By improving diagnostic yield and mechanistic insight, OGM has the potential to inform patient management and refine genotype–phenotype correlations in neurological disorders of somatic etiology. In the case of our patient with epilepsy and cortical dysplasia, the identification of the somatic second hit by OGM was key to explaining the patient’s full clinical presentation. Understanding these disease mechanisms, such as the phenotypic associations of single vs. bi-allelic gene inactivation, may lead to improved treatment approaches in the future.

## Methods

### Brain tissue

One C57BL6 mouse was humanely euthanized under deep anesthesia, and then brain tissue was rapidly dissected, frozen on dry ice, and stored at -80°C. Experiments were performed in compliance with the animal care guidelines issued by the National Institutes of Health and the institutional animal care and use committee (AR20-00080) at Nationwide Children’s Hospital.

Patients undergoing neurosurgical treatment for drug-resistant epilepsy at Nationwide Children’s Hospital were enrolled via written consent on an Institutional Review Board (IRB)-approved research protocol (IRB18-00786). Surgically resected brain tissue was labeled by anatomic region and divided for clinical neuropathologic evaluation and for genomic analysis. Samples for genomic analysis were stored frozen at -80°C immediately following surgical resection.

### DNA isolation

High-Molecular Weight (HMW) genomic DNA (gDNA) was isolated from human brain tissue according to the *Bionano Prep SP Brain Tissue DNA Isolation Tech Note* (Document number 30400, Revision A; Bionano Genomics). Briefly, 15 mg of frozen brain tissue was used per sample. Tissue was homogenized and filtered, followed by pelleting and sequential enzymatic digestion. DNA was bound to a Nanobind disk, washed to remove impurities, and eluted in buffer. The recovered HMW gDNA was then gently homogenized and equilibrated at room temperature to improve sample uniformity before quantification.

### Optical genome mapping

HMW gDNA was labeled using the Bionano Prep DLS-G2 Labeling kit (Bionano Genomics) following the manufacturer’s protocol. A total of 750 ng DNA per sample was enzymatically labeled with the DL-Green fluorophore using DLE-1. Excess label was removed, after which the DNA was stained and quantified using the Qubit dsDNA HS kit (Thermo Fisher). Labeled and stained gDNA was loaded onto a G3.3 Saphyr Chip according to the manufacturer’s *Saphyr System User Guide* with a target of 1600 Gb data collection. All samples achieved 1600 Gb except Patient 1 which reached 882 Gb collected.

### PacBio HiFi Sequencing

High molecular weight genomic DNA was extracted from brain tissue using the Bionano Brain protocol and sheared using a Megaruptor 3 (1000 ng input per sample; speed setting 29-32; 3-5 passes depending on viscosity), generating mean fragment sizes ranging from 28.5 kb to 32.5 kb. Between 750 ng and 950 ng of sheared DNA was carried forward into library preparation using the SMRTbell Express Template Prep Kit 3.0 (PacBio) with the following modifications: DNA damage repair, end-repair, and A-tailing incubations were extended to 1 hr, and adapter ligation was performed overnight. Incompletely formed SMRTbell molecules were removed by exonuclease treatment, which was extended to 45 min. Final libraries were size selected on a BluePippin instrument (Sage Science) using a >8.5 kb cutoff cassette. The size-selected SMRTbell libraries were sequenced on the PacBio Revio system using SPRQ chemistry at an on-plate loading concentration ranging between 101 pM and 165 pM. Each library was sequenced on a single 25M SMRT cell using a 30 hr movie collection time. Circular consensus sequencing (CCS) analysis was performed using SMRT Link v25.2.0.266456 with default parameters to generate HiFi reads. Across all four libraries, HiFi yields ranged from 84.7 Gb to 130.5 Gb (5.1 - 8.3 million reads) with mean HiFi lengths of 15.7 kb - 18.4 kb. Secondary analysis was carried out using the “Variant Calling” application in SMRT Link with default parameters, which generated variant call format (VCF) files and aligned BAM files against the GRCh38/hg38 reference genome. Mean mapped coverage per sample ranged from 27x to 41x.

The probability of at least 5 reads identifying the variant at the VAF called by OGM given the mean mapped coverage of each sample was calculated using a binomial distribution.

### Amplicon validation

Long-range PCR was performed using PrimeSTAR LongSeq DNA Polymerase (Takara Bio, Cat. #R045A) according to the manufacturer’s recommendations. PCR reactions were set up with 100 ng genomic DNA input from either NA19240 control DNA (Coriell) or Patient 2 brain DNA (sheared PacBio input). Primer sequences used to amplify the region spanning the putative DEPDC5 deletion were: Forward: 5′-CTTTTGACAGCTGCTACTGGCTTGTCG-3 and Reverse: 5′-GGATACCGTGAACAGTAACTCCCTAGC-3′. Thermocycling was carried out using the following program: initial denaturation at 94 °C for 1 min; 30 cycles of 98 °C for 10–20 s, 55 °C for 15 s, and 68 °C for 9 min; followed by a final hold at 4 °C. PCR products were purified using a 1x SPRIselect bead cleanup (Beckman Coulter) and eluted in 25 µL of EB. Amplicons were visualized on an Agilent Genomic DNA ScreenTape using the Agilent TapeStation 4200 system, which resolved the expected reference (∼14 kb) and deletion (850 bp) products. To enrich the deletion product prior to Sanger sequencing, PCR amplicons were subjected to bead-based size selection. A diluted AMPure PB bead mixture was prepared by combining 130 µL of EB with 70 µL of AMPure PB beads. A 3.1x diluted AMPure PB cleanup was then performed using 35% AMPure PB beads (185 µL diluted beads added to 25 µL PCR product), corresponding to an approximate 5 kb cutoff. This approach preferentially retained the ∼850 bp deletion product while depleting the ∼14 kb reference amplicon. A subsequent QIAquick PCR purification (Qiagen) further reduced residual high–molecular weight background. Sanger sequencing was carried out using the Applied Biosystems BigDye Terminator v3.1 Cycle Sequencing Kit, following the manufacturer’s protocol. Sequencing reactions were analyzed by capillary electrophoresis on an ABI Genetic Analyzer. Chromatogram files were processed and interpreted using Sequencher (Gene Codes) and Mutation Surveyor (SoftGenetics) with default parameters, which confirmed the breakpoint junctions of the *DEPDC5* deletion.

### Data analysis

For optical genome mapping, analysis was performed using the Rare Variant Analysis pipeline in Bionano Access 1.8.2.1 and Bionano Solve 3.8.2.1 to identify somatic structural variants. Molecules were aligned to the GRCh38 human reference genome and structural variant calls were annotated with overlapping or nearest non-overlapping gene, variant allele fraction, and population frequency based on a database of 285 control samples provided by Bionano. Variants were filtered for <40% variant allele fraction to capture likely somatic variants and for population frequency <10% for rare somatic variant detection or 0% for potential disease-causing variants.

### Structural Variant Annotation

To characterize the genomic context of the reference regions containing each structural variant, all variants were examined using the UCSC Genome Browser with hg38 human genome assembly (Spyrou et al. 2024). Genic and intergenic regions were identified using the NCBI RefSeq Genes (curated subset) track. Regions overlapping a RefSeq gene were classified as *intragenic*, whereas regions without overlap were classified as *intergenic*.

Regulatory elements were also assessed using the ENCODE3 Registry of Candidate Cis-Regulatory Elements (cCREs) track. We quantified the number of enhancer-like, including both proximal and distal, and promoter-like signatures for each region. Regions containing ≥ 5 enhancer-like signatures or any promoter-like signatures were considered enriched for those respective features. To assess repetitive element content, the reference start and end positions of each structural variant were extracted and converted to FASTA sequences using bedtools2 (Quinlan and Hall 2010). These sequences were then annotated using RepeatMasker to determine the proportion of each reference region composed of repetitive elements, including short interspersed nuclear elements (SINEs) and long interspersed nuclear elements (LINEs) (Chen 2004). Regions containing ≥ 2 fold percent composition of repetitive elements were considered likely enriched for these elements, based on the approximate genome-wide content of 13% SINEs and 21% LINEs in the human genome (Zhang et al. 2021). To assess whether these variants were significantly enriched for repetitive elements, we performed permutation testing using regioneR (Gel et al. 2016). For each reference region containing a structural variant called by OGM, 10,000 random intervals of equivalent length were sampled from the mappable portion of the same chromosome to account for local genomic architecture and ensure a biologically relevant null distribution. Statistical significance was determined by comparing the total base-pair overlap of the observed reference region to the distribution of overlaps of these permuted regions.

To assess potential disease relevance, we first filtered our list of overlap or nearest non overlap genes for protein coding genes using Ensembl’s BioMart tool (Dyer et al. 2025). We then cross-referenced this list with curated gene sets including epilepsy associated genes and neurodevelopmental disorder genes from the SysNDD database, including definitive, moderate and limited evidence categories (Kochinke et al. 2016; Bedrosian et al. 2022). Additionally, all overlap genes were queried in OMIM, noting associations with neurological phenotypes. Finally, the reference regions containing the structural variants were annotated using AnnotSV (Geoffroy et al. 2018).

To assess the frequency of the variants called by OGM in the population we used gnomAD SV v.4.1.0 and queried the reference start and end regions called by Bionano’s analysis software (Collins et al. 2020). We manually reviewed and counted the number of variants of similar structural variant type, noting both the range of size and allele count. Given that OGM resolution is limited by label sequence and density, insertions called by OGM are often identified as duplications when base pair resolution is achieved. Therefore, for all insertion events called by OGM, we noted both insertion and duplication occurrence observed in gnomAD SV.

## Data Access

The data generated in this study have been deposited in Synapse under project accession syn74372113.

## Data Availability

All data produced are available online at Synapse under project accession syn74372113.

## Acknowledgements

Funding was provided by NIH-NINDS (R01NS129784) to T.A.B. The content is solely the responsibility of the authors and does not necessarily represent the official views of the NIH. Some figures were created in BioRender. Yassmine Akkari for editing and feedback. AM, KEM, and TAB conceptualized the study. AD, JL, JP, AS, CP, DB, DLT, APO, and ERM contributed to patient consent, acquisition of clinical samples, clinical phenotyping, and/or variant interpretation. MEHG, LRV, ES, and LMW performed the experiments. AM, JJA, DK, and TAB analyzed the data. AM, JJA, KEM, and TAB wrote the manuscript. All authors reviewed and edited the manuscript.

## References

1. Ade C, Roy-Engel AM, Deininger PL. 2013. Alu elements: an intrinsic source of human genome instability. Curr Opin Virol 3: 639–645.

2. Bedrosian TA, Miller KE, Grischow OE, Schieffer KM, LaHaye S, Yoon H, Miller AR, Navarro J, Westfall J, Leraas K et al. 2022. Detection of brain somatic variation in epilepsy-associated developmental lesions. Epilepsia 63: 1981–1997.

3. Blumcke I, Spreafico R, Haaker G, Coras R, Kobow K, Bien CG, Pfafflin M, Elger C, Widman G, Schramm J et al. 2017. Histopathological Findings in Brain Tissue Obtained during Epilepsy Surgery. N Engl J Med 377: 1648–1656.

4. Chen N. 2004. Using RepeatMasker to identify repetitive elements in genomic sequences. Curr Protoc Bioinformatics **Chapter 4**: Unit 4 10.

5. Chung C, Yang X, Bae T, Vong KI, Mittal S, Donkels C, Westley Phillips H, Li Z, Marsh APL, Breuss MW et al. 2023. Comprehensive multi-omic profiling of somatic mutations in malformations of cortical development. Nat Genet 55: 209–220.

6. Collins RL, Brand H, Karczewski KJ, Zhao X, Alfoldi J, Francioli LC, Khera AV, Lowther C, Gauthier LD, Wang H et al. 2020. A structural variation reference for medical and population genetics. Nature 581: 444–451.

7. Corrigan RR, Mashburn-Warren LM, Yoon H, Bedrosian TA. 2025. Somatic Mosaicism in Brain Disorders. Annu Rev Pathol 20: 13–32.

8. Deng R, Perenthaler E, Nikoncuk A, Yousefi S, Lanko K, Schot R, Maresca M, Medico-Salsench E, Sanderson LE, Parker MJ et al. 2024. BRAIN-MAGNET: A novel functional genomics atlas coupled with convolutional neural networks facilitates clinical interpretation of disease relevant variants in non-coding regulatory elements. medRxiv doi:10.1101/2024.04.13.24305761: 2024.2004.2013.24305761.

9. Dyer SC, Austine-Orimoloye O, Azov AG, Barba M, Barnes I, Barrera-Enriquez VP, Becker A, Bennett R, Beracochea M, Berry A et al. 2025. Ensembl 2025. Nucleic Acids Res 53: D948–D957.

10. Gel B, Diez-Villanueva A, Serra E, Buschbeck M, Peinado MA, Malinverni R. 2016. regioneR: an R/Bioconductor package for the association analysis of genomic regions based on permutation tests. Bioinformatics 32: 289–291.

11. Geoffroy V, Herenger Y, Kress A, Stoetzel C, Piton A, Dollfus H, Muller J. 2018. AnnotSV: an integrated tool for structural variations annotation. Bioinformatics 34: 3572–3574.

12. Kobow K, Jabari S, Pieper T, Kudernatsch M, Polster T, Woermann FG, Kalbhenn T, Hamer H, Rossler K, Muhlebner A et al. 2020. Mosaic trisomy of chromosome 1q in human brain tissue associates with unilateral polymicrogyria, very early-onset focal epilepsy, and severe developmental delay. Acta Neuropathol 140: 881–891.

13. Kochinke K, Zweier C, Nijhof B, Fenckova M, Cizek P, Honti F, Keerthikumar S, Oortveld MA, Kleefstra T, Kramer JM et al. 2016. Systematic Phenomics Analysis Deconvolutes Genes Mutated in Intellectual Disability into Biologically Coherent Modules. Am J Hum Genet 98: 149–164.

14. Lai D, Gade M, Yang E, Koh HY, Lu J, Walley NM, Buckley AF, Sands TT, Akman CI, Mikati MA et al. 2022. Somatic variants in diverse genes leads to a spectrum of focal cortical malformations. Brain 145: 2704–2720.

15. Leng K, Cadwell CR, Devine WP, Tihan T, Qi Z, Singhal NS, Glenn OA, Kamiya S, Wiita AP, Berger AC et al. 2024. Cell-Type Specificity of Mosaic Chromosome 1q Gain Resolved by snRNA-seq in a Case of Epilepsy With Hyaline Protoplasmic Astrocytopathy. Neurol Genet 10: e200142.

16. Levy B, Burnside RD, Akkari Y. 2025. Optical Genome Mapping: A New Tool for Cytogenomic Analysis. Genes (Basel) 16.

17. Lopez-Rivera JA, Leu C, Macnee M, Khoury J, Hoffmann L, Coras R, Kobow K, Bhattarai N, Perez-Palma E, Hamer H et al. 2023. The genomic landscape across 474 surgically accessible epileptogenic human brain lesions. Brain 146: 1342–1356.

18. McConnell MJ, Lindberg MR, Brennand KJ, Piper JC, Voet T, Cowing-Zitron C, Shumilina S, Lasken RS, Vermeesch JR, Hall IM et al. 2013. Mosaic copy number variation in human neurons. Science 342: 632–637.

19. Miller KE, Rivaldi AC, Shinagawa N, Sran S, Navarro JB, Westfall JJ, Miller AR, Roberts RD, Akkari Y, Supinger R et al. 2023. Post-zygotic rescue of meiotic errors causes brain mosaicism and focal epilepsy. Nat Genet 55: 1920–1928.

20. Neveling K, Mantere T, Vermeulen S, Oorsprong M, van Beek R, Kater-Baats E, Pauper M, van der Zande G, Smeets D, Weghuis DO et al. 2021. Next-generation cytogenetics: Comprehensive assessment of 52 hematological malignancy genomes by optical genome mapping. Am J Hum Genet 108: 1423–1435.

21. Pang AWC, Kosco K, Sahajpal NS, Sridhar A, Hauenstein J, Clifford B, Estabrook J, Chitsazan AD, Sahoo T, Iqbal A et al. 2023. Analytic Validation of Optical Genome Mapping in Hematological Malignancies. Biomedicines 11.

22. Quinlan AR, Hall IM. 2010. BEDTools: a flexible suite of utilities for comparing genomic features. Bioinformatics 26: 841–842.

23. Rehen SK, McConnell MJ, Kaushal D, Kingsbury MA, Yang AH, Chun J. 2001. Chromosomal variation in neurons of the developing and adult mammalian nervous system. Proc Natl Acad Sci U S A 98: 13361–13366.

24. Ribierre T, Deleuze C, Bacq A, Baldassari S, Marsan E, Chipaux M, Muraca G, Roussel D, Navarro V, Leguern E et al. 2018. Second-hit mosaic mutation in mTORC1 repressor DEPDC5 causes focal cortical dysplasia-associated epilepsy. J Clin Invest 128: 2452–2458.

25. Richards S, Aziz N, Bale S, Bick D, Das S, Gastier-Foster J, Grody WW, Hegde M, Lyon E, Spector E et al. 2015. Standards and guidelines for the interpretation of sequence variants: a joint consensus recommendation of the American College of Medical Genetics and Genomics and the Association for Molecular Pathology. Genet Med 17: 405–424.

26. Rohrback S, Siddoway B, Liu CS, Chun J. 2018. Genomic mosaicism in the developing and adult brain. Dev Neurobiol 78: 1026–1048.

27. Sahajpal NS, Barseghyan H, Kolhe R, Hastie A, Chaubey A. 2021. Optical Genome Mapping as a Next-Generation Cytogenomic Tool for Detection of Structural and Copy Number Variations for Prenatal Genomic Analyses. Genes (Basel) 12.

28. Sahajpal NS, Mondal AK, Singh H, Vashisht A, Ananth S, Saul D, Hastie AR, Hilton B, DuPont BR, Savage NM et al. 2023. Clinical Utility of Optical Genome Mapping and 523-Gene Next Generation Sequencing Panel for Comprehensive Evaluation of Myeloid Cancers. Cancers (Basel) 15.

29. Sekar S, Tomasini L, Proukakis C, Bae T, Manlove L, Jang Y, Scuderi S, Zhou B, Kalyva M, Amiri A et al. 2020. Complex mosaic structural variations in human fetal brains. Genome Res 30: 1695–1704.

30. Spyrou J, Aung KP, Vanyai H, Leventer RJ, Maljevic S, Lockhart PJ, Howell KB, Reid CA. 2024. Slc35a2 mosaic knockout impacts cortical development, dendritic arborisation, and neuronal firing. Neurobiol Dis 201: 106657.

31. Sran S, Ringland A, Bedrosian TA. 2024. Building the brain mosaic: an expanded view. Trends Genet 40: 747–756.

32. Vasudevaraja V, Rodriguez JH, Pelorosso C, Zhu K, Buccoliero AM, Onozato M, Mohamed H, Serrano J, Tredwin L, Garonzi M et al. 2021. Somatic Focal Copy Number Gains of Noncoding Regions of Receptor Tyrosine Kinase Genes in Treatment-Resistant Epilepsy. J Neuropathol Exp Neurol 80: 160–168.

33. Zhang XO, Pratt H, Weng Z. 2021. Investigating the Potential Roles of SINEs in the Human Genome. Annu Rev Genomics Hum Genet 22: 199–218.

