## Supplemental Material for "Optical genome mapping identifies clinically relevant somatic structural variation in epilepsy-affected brain tissue"

**This PDF file includes:**

Supplementary Table S1

Supplementary Figure S1

| Patient ID | Chromosome | RefStartPos (bp) | RefEndPos (bp) | Type | Size (bp) | ISCN | Overlap Genes |
|---|---|---|---|---|---|---|---|
| Patient 1 | 5 | 14993191 | 15008321 | insertion | 5,939 | *gm[GRCh38] ins(5;?)(p15.2p15.1;?)(14993191_15008321;?* | - |
| Patient 1 | 7 | 65671676 | 65672645 | insertion | 27,428 | *ogm[GRCh38] ins(7;?)(q11.21;?)(65671676_65672645;?)* | *LINC03006;INTS4P2* |
| Patient 1 | 7 | 152306940 | 152327378 | deletion | 5,045 | *ogm[GRCh38] 7q36.1(152306940_152327378)x1* | *KMT2C* |
| Patient 1 | 9 | 63391388 | 63401812 | insertion | 5,927 | *ogm[GRCh38] ins(9;?)(q13;?)(63391388_63401812;?)* | - |
| Patient 1 | 10 | 5359177 | 5374850 | insertion | 10,215 | *ogm[GRCh38] ins(10;?)(p15.1;?)(5359177_5374850;?)* | *UCN3* |
| Patient 1 | 19 | 40849294 | 40891686 | deletion | 31,764 | *ogm[GRCh38] 19q13.2(40849294_40891686)x1* | *CYP2A6;CYP2A7;CYP2G1P* |
| Patient 1 | 22 | 42113933 | 42139458 | deletion | 12,136 | *ogm[GRCh38] 22q13.2(42113933_42139458)x1* | *NDUFA6-DT;CYP2D6* |
| Patient 2 | 1 | 120434755 | 120475182 | insertion | 6,504 | *ogm[GRCh38] ins(1;?)(p11.2;?)(120434755_120475182;?)* | *NBPF8;PDE4DIPP2* |
| Patient 2 | 1 | 208265618 | 208273500 | insertion | 13,368 | *ogm[GRCh38] ins(1;?)(q32.2;?)(208265618_208273500;?)* | - |
| Patient 2 | 6 | 54980641 | 55004387 | deletion | 5,304 | *ogm[GRCh38] 6p12.1(54980641_55004387)x1* | - |
| Patient 2 | 11 | 4289557 | 4340719 | duplication | 51,162 | *ogm[GRCh38] dup(11)(p15.4p15.4)(4289557_4340719)* | *SSU72L3;SSU72L1* |
| Patient 2 | 15 | 23038484 | 23057512 | insertion | 22,318 | *ogm[GRCh38] ins(15;?)(q11.2;?)(23038484_23057512;?)* | *TUBGCP5* |
| Patient 2 | 16 | 33738145 | 33791415 | deletion | 5,497 | *ogm[GRCh38] 16p11.2(33738145_33791415)x1* | *ENPP7P13* |
| Patient 2 | 17 | 15198788 | 15297128 | deletion | 14,103 | *ogm[GRCh38] 17p12(15198788_15297128)x1* | *PMP22;MIR4731* |
| Patient 2 | 18 | 15195003 | 15218872 | deletion | 5,171 | *ogm[GRCh38] 18p11.21(15195003_15218872)x1* | - |
| Patient 2 | 22 | 31752788 | 31774605 | deletion | 11,876 | *ogm[GRCh38] 22q12.2(31752788_31774605)x1* | *DEPDC5* |
| Patient 3 | 1 | 9266521 | 9339579 | duplication | 73,058 | *ogm[GRCh38] dup(1)(p36.22p36.22)(9266521_9339579)* | *H6PD;SPSB1* |
| Patient 3 | 3 | 6599252 | 6610335 | deletion | 5,386 | *ogm[GRCh38] 3p26.1(6599252_6610335)x1* | *LOC105376944* |
| Patient 3 | 4 | 160670006 | 160694101 | insertion | 10,872 | *ogm[GRCh38] ins(4;?)(q32.1;?)(160670006_160694101;?)* | - |
| Patient 3 | 6 | 142104479 | 142155808 | deletion | 30,304 | *ogm[GRCh38] 6q24.1(142104479_142155808)x1* | *NMBR;GJE1;VTA1* |
| Patient 3 | 9 | 105032351 | 105056294 | deletion | 5,293 | *ogm[GRCh38] 9q31.1(105032351_105056294)x1* | - |
| Patient 3 | 10 | 87214661 | 87350880 | deletion | 131,604 | *ogm[GRCh38] 10q23.2(87214661_87350880)x1* | *NUTM2A;NUTM2A-AS1;LINC00863* |
| Patient 3 | 12 | 8209061 | 8211161 | insertion | 29,612 | *ogm[GRCh38] ins(12;?)(p13.31;?)(8209061_8211161;?)* | - |
| Patient 3 | 12 | 79755184 | 79778647 | insertion | 23,264 | *ogm[GRCh38] ins(12;?)(q21.2;?)(79755184_79778647;?)* | *PPP1R12A-AS2;PPP1R12A* |
| Patient 3 | 13 | 98596625 | 98638935 | deletion | 5,571 | *ogm[GRCh38] 13q32.2(98596625_98638935)x1* | - |
| Patient 3 | 16 | 14685557 | 14776527 | deletion | 78,164 | *ogm[GRCh38] 16p13.12p13.11(14685557_14776527)x1* | *PLA2G10;NPIPA3;NPIPA2* |
| Patient 3 | 16 | 29061167 | 29463027 | inversion | 366,830 | *ogm[GRCh38] inv(16)(p11.2p11.2)(29039401_29498408)* | *X29P2; NPIPB11; SMG1P6; BOLA2-SMG1P6; LOC606724; BOLA2; SLX1B; SLX1B-SULT1A4; SULT1A4; LOC388242; NPIPB12; RRN3P2;LOC101928188;LOC1027237* |
| Patient 3 | 17 | 82339478 | 82365763 | insertion | 17,938 | *ogm[GRCh38] ins(17;?)(q25.3;?)(82339478_82365763;?)* | *TEX19* |
| Patient 4 | 1 | 149345419 | 149599348 | duplication | 248,591 | *ogm[GRCh38] dup(1)(q21.2q21.2)(149345419_149594010)* | *SEC22B2P;NOTCH2NLC;NBPF19;PPIAL4C* |
| Patient 4 | 2 | 183921082 | 183939473 | deletion | 9,689 | *ogm[GRCh38] 2q32.1(183921082_183939473)x1* | - |
| Patient 4 | 6 | 167933101 | 168197312 | duplication | 264,212 | *ogm[GRCh38] dup(6)(q27q27)(167933101_168197312)* | *AFDN;HGC6.3;KIF25-AS1;KIF25;FRMD1* |
| Patient 4 | 7 | 96911252 | 96928401 | deletion | 3,253 | *ogm[GRCh38] 7q21.3(96911252_96928401)x1* | - |
| Patient 4 | 8 | 8010904 | 8060871 | insertion | 23,801 | *ogm[GRCh38] ins(8;?)(p23.1;?)(8010904_8060871;?)* | *FAM90A11;FAM90A24;FAM90A12* |
| Patient 4 | 17 | 37975673 | 38032811 | insertion | 22,475 | *ogm[GRCh38] ins(17;?)(q12;?)(37975673_38032811;?)* | *TBC1D3L;TBC1D3D* |

| Nearest NonOverlap Gene | Putative Gene Fusion | VAF | VAF Supportive of Somatic Origin | Confidence | Frequency in Control Database | Number of Similar SV Type Variants in gnomAD SV | gnomAD SV Size Range | gnomAD SV Allele Count Range | Number of Enhancer-like Elements (ENCODE3 cCREs) | Number of Promoter-like Elements (ENCODE3 cCREs) | Percent Overlap SINEs (RepeatMasker) | SINE Permutation Test p value |
|---|---|---|---|---|---|---|---|---|---|---|---|---|
| *ANKH* | - | 0.4 | Ambiguous | 0.98 | 0.7 | 11 DUP; 1 INS | 227 bp - 1.30Mb; 58bp | 1-884; 5 | 4 | 0 | 19.89 | 0.153484652 |
| *CCT6P1* | - | 0.3 | Suggestive | 0.99 | 0.4 | 3 DUP | 70-689 kb | 87-677 | 0 | 0 | 0 | 1 |
| *FABP5P3* | - | 0.4 | Ambiguous | 0.99 | 0.4 | 9 DEL | 85 bp - 451 kb | 1-1128 | 7 | 0 | 24.32 | 0.154284572 |
| *LOC100996643* | - | 0.4 | Ambiguous | 0.98 | 7 | 4 DUP | 6 kb - 418 kb | 26-2,302 | 7 | 2 | 6.86 | 0.668433157 |
| *TUBAL3* | - | 0.4 | Ambiguous | 0.99 | 0 | 31 DUP; 3 INS | 56 bp - 2.58 Mb; 50-59 bp | 1-7,029; 5-3,918 | 6 | 1 | 10.53 | 0.709729027 |
| *CYP2B7P* | *CYP2A6-CYP2G1P* | 0.4 | Ambiguous | 0.99 | 5.3 | 12 DEL | 55 bp - 31 kb | 1-38,797 | 8 | 0 | 28.64 | 0.488751125 |
| *CYP2D7* | - | 0.4 | Ambiguous | 0.99 | 7.4 | 7 DEL | 105 bp - 18.4 kb | 1-609 | 6 | 1 | 17.02 | 0.443755624 |
| *PFN1P2* | - | 0.4 | Suggestive | 0.98 | 4.2 | 12 DUP; 2 INS | 5 kb - 6.57 Mb; 280 bp | 2-905; 1-2 | 0 | 0 | 14.29 | 0.530646935 |
| *PLXNA2* | - | 0.4 | Ambiguous | 0.99 | 0 | 1 DUP | 13 kb | 532 | 11 | 0 | 10.18 | 0.547045295 |
| *FAM83B* | - | 0.4 | Suggestive | 0.99 | 9.1 | 6 DEL | 651 bp - 14.4 kb | 1-107,679 | 5 | 0 | 10.16 | 0.452054795 |
| *SSU72L4* | - | 0.2 | Strongly | 0.99 | 3.2 | 9 DUP | 7.5-355 kb | 1-19,109 | 0 | 0 | 7.96 | 0.767823218 |
| *CYFIP1* | - | 0.4 | Ambiguous | 0.99 | 9.1 | 125 DUP; 2 INS | 56 bp - 5.35 Mb; 106-280 bp | 1-39,500; 2-3 | 5 | 1 | 9.22 | 0.561843816 |
| *TP53TG3F* | - | 0.3 | Suggestive | 0.99 | 1.4 | 12 DEL | 70 bp - 172 kb | 1-12,543 | 0 | 0 | 8 | 0.817218278 |
| *TEKT3* | - | 0.3 | Suggestive | 0.99 | 0.4 | 22 DEL | 58 bp - 1.33 Mb | 1-4,985 | 47 | 5 | 10.39 | 0.860613939 |
| *LOC644669* | - | 0.3 | Suggestive | 0.99 | 1.4 | 9 DEL | 76 bp - 31 kb | 1-814 | 0 | 0 | 5.71 | 0.755924408 |
| *PRR14L* | - | 0.2 | Strongly | 0.99 | 0 | 5 DEL | 95 bp - 1.92 kb | 1-40 | 6 | 3 | 55.51 | 0.00679932 |
| *LNCTAM34A* | *H6PD-SPSB1* | 0.3 | Suggestive | 0.99 | 0.7 | 13 DUP | 51 bp - 418 kb | 1-307 | 56 | 1 | 17.6 | 0.275072493 |
| *GRM7-AS3* | - | 0.4 | Ambiguous | 0.99 | 8.8 | 5 DEL | 4.49 kb - 8.60 Mb | 1-13,716 | 0 | 0 | 7.98 | 0.618338166 |
| *FSTL5* | - | 0.2 | Suggestive | 0.99 | 1.1 | 10 DUP; 6 INS | 55 bp - 836 kb; 56-534 bp | 1-28,919; 1-374 | 0 | 0 | 8.52 | 0.490850915 |
| *ADGRG6* | *NMBR-VTA1* | 0.1 | Strongly | 0.99 | 0 | 11 DEL | 88 bp - 92.5 kb | 1-6 | 5 | 1 | 7 | 0.687731227 |
| *CT70* | - | 0.3 | Suggestive | 0.99 | 1.4 | 8 DEL | 52 bp- 8.63 kb | 1-15,192 | 13 | 0 | 22.55 | 0.129587041 |
| *NUTM2D* | - | 0.4 | Ambiguous | 0.98 | 7.7 | 20 DEL | 57 bp - 129 kb | 1-12,819 | 4 | 1 | 10.74 | 0.554344566 |
| *FAM66C* | - | 0.1 | Strongly | 0.99 | 2.8 | 3 DUP; 1 INS | 14.3-966 kb; 281 bp | 2-19,152; 3 | 0 | 0 | 13.38 | 0.399260074 |
| *PAWR* | - | 0.4 | Ambiguous | 0.99 | 3.9 | 5 DUP; 1 INS | 61 bp - 190 kb; 265 bp | 1-7; 2 | 8 | 0 | 12.73 | 0.440855914 |
| *STK24-AS1* | - | 0.4 | Ambiguous | 0.99 | 6 | 6 DEL | 64 bp - 6.78 kb | 1- 249 | 10 | 0 | 36.27 | 0.00639936 |
| *BFAR* | - | 0.2 | Strongly | 0.99 | 9.1 | 9 DEL | 81 bp - 4.03 Mb | 1-18,177 | 1 | 0 | 43.82 | 0.0189981 |
| *LAT* | - | 0.3 | Suggestive | 0.98 | 0.4 | 1 INV | 15.2 Mb | 2 | 132 | 6 | 20.95 | 0.386261374 |
| *SECTM1* | - | 0.4 | Ambiguous | 0.99 | 4.2 | 33 DUP; 7 INS | 51 bp - 887 kb; 50-281 bp | 1- 33,036; 1-6,144 | 22 | 1 | 14.92 | 0.648335166 |
| *LINC00869* | - | 0.1 | Strongly | 0.97 | 1.8 | 29 DUP | 6 kb - 9.12 Mb | 1-16,893 | 1 | 1 | 9.81 | 0.641635836 |
| *MIR548AE1* | - | 0.4 | Ambiguous | 0.93 | 5.3 | 7 DEL | 91 bp - 173 kb | 1-4,040 | 2 | 0 | 1.6 | 0.96450355 |
| *LOC105378137* | - | 0.2 | Strongly | 0.97 | 1.4 | 65 DUP | 50 bp - 744 kb | 1-57,014 | 120 | 3 | 6.16 | 0.851214879 |
| *DLX6-AS1* | - | 0.3 | Suggestive | 0.98 | 6 | 4 DEL | 143 bp - 6.58 kb | 1-34,453 | 8 | 0 | 11.13 | 0.472952705 |
| *FAM66E* | - | 0.3 | Suggestive | 0.99 | 1.8 | 7 DUP; 2 INS | 7.19-307 kb; 279 bp | 41-25,625; 1-2 | 0 | 0 | 8.2 | 0.639336066 |
| *TBC1D3C* | - | 0.2 | Suggestive | 0.99 | 8.4 | 2 DUP; 1 INS | 11.8-12 kb; 54 bp | 5,079-8,792; 108 | 0 | 0 | 18.17 | 0.528647135 |

| SINE Permutation Test z score | Percent Overlap LINEs (RepeatMasker) | LINE Permutation Test p value | LINE Permutation Test z score | Curated Lists and OMIM ID | OMIM Neurological Phenotype | AnnotSV ACMG Score of OGM Reference Regions | Support Found in PacBio | PacBio Coverage | Probabilty of ≥5 Reads in PacBio | Fit for Patient Phenotype |
|---|---|---|---|---|---|---|---|---|---|---|
| 0.8481 | 18.29 | 0.698630137 | -0.661 | All, OMIM:605145 | No | - | Yes | 27x | 0.99 | No |
| -0.7032 | 24.05 | 0.370162984 | -0.2054 | - | - | - | No | 27x | 0.95 | - |
| 0.9268 | 27.48 | 0.342365763 | 0.2151 | All, OMIM:606833 | Yes | Likely Pathogenic (4) | Yes | 27x | 0.99 | No |
| -0.5599 | 0 | 1 | -0.9952 | - | - | - | No | 27x | 0.99 | - |
| -0.6153 | 14.18 | 0.602339766 | -0.4787 | SysNDD | - | - | Yes | 27x | 1.00 | No |
| 0.0756 | 15.3 | 0.358164184 | 0.099 | OMIM:122720 | No | Pathogenic (5) | Yes | 27x | 1.00 | No |
| 0.0649 | 22.01 | 0.176482352 | 0.8691 | Epilepsy, OMIM:124030 | No | Variant of Uncertain Signifcance (3) | Yes | 27x | 0.99 | No |
| -0.2933 | 21.61 | 0.446455354 | -0.0313 | - | - | - | No | 41x | 1.00 | - |
| -0.3307 | 5.84 | 0.788821118 | -0.8183 | Epilepsy and SysNDD | - | - | Yes | 41x | 1.00 | No |
| -0.1774 | 32.86 | 0.280171983 | 0.4678 | - | - | Variant of Uncertain Signifcance (3) | Yes | 41x | 1.00 | - |
| -0.693 | 41.57 | 0.121487851 | 1.2164 | - | - | Benign (1) | No | 41x | 0.93 | - |
| -0.2764 | 22.17 | 0.313968603 | 0.2772 | SysNDD | - | - | Yes | 41x | 1.00 | No |
| -0.868 | 16.48 | 0.430856914 | -0.0102 | - | - | - | No | 41x | 1.00 | - |
| -1.0456 | 29.03 | 0.111688831 | 1.2369 | OMIM:601097 | Yes | Pathogenic (5) | Yes | 41x | 1.00 | No |
| -0.6246 | 41.37 | 0.124587541 | 1.23 | - | - | Benign (1) | Yes | 41x | 1.00 | - |
| 2.6058 | 5.08 | 0.628137186 | -0.5535 | All, OMIM:614191 | Yes | Pathogenic (5) | Yes | 41x | 0.93 | Yes |
| 0.3474 | 8.58 | 0.875612439 | -1.0529 | OMIM:138090 |  | Variant of Uncertain Signifcance (3) | Yes | 34x | 1.00 | No |
| -0.4443 | 52.07 | 0.105189481 | 1.4401 | - | - | Benign (1) | Yes | 34x | 1.00 | - |
| -0.2191 | 55.72 | 0.061093891 | 1.7785 | - | - | - | No | 34x | 0.90 | - |
| -0.5779 | 33.12 | 0.256674333 | 0.5905 | SysNDD, OMIM:612243 | No | Variant of Uncertain Signifcance (3) | No | 34x | 0.09 | No |
| 1.1042 | 27.57 | 0.310668933 | 0.3568 | - | - | Variant of Uncertain Signifcance (3) | Yes | 34x | 0.96 | - |
| -0.3912 | 26.35 | 0.291970803 | 0.3905 | - | - | Benign (1) | No | 34x | 1.00 | - |
| -0.0281 | 4.9 | 1 | -0.7687 | - | - | - | Yes | 34x | 0.39 | - |
| -0.1885 | 38.87 | 0.173882612 | 0.9205 | SysNDD, OMIM:602021 | Yes | - | Yes | 34x | 1.00 | No |
| 4.0056 | 27.59 | 0.297170283 | 0.5093 | - | - | Variant of Uncertain Signifcance (3) | Yes | 34x | 1.00 | - |
| 2.1371 | 7.32 | 0.814418558 | -0.8666 | - | - | Variant of Uncertain Signifcance (3) | Yes | 34x | 0.80 | - |
| 0.2621 | 14.87 | 0.586841316 | -0.1937 | - | - | - | No | 34x | 0.99 | - |
| -0.4945 | 3.85 | 0.885611439 | -0.987 | - | - | - | Yes | 34x | 1.00 | - |
| -0.5013 | 17.12 | 0.733626637 | -0.6105 | OMIM:618025 | Yes | Variant of Uncertain Signifcance (3) | No | 31x | 0.25 | No |
| -1.2037 | 38.24 | 0.180581942 | 0.9127 | - | - | Variant of Uncertain Signifcance (3) | Yes | 31x | 1.00 | - |
| -0.8379 | 13.47 | 0.897010299 | -1.1987 | Epilepsy | - | Benign (1) | Yes | 31x | 0.57 | No |
| -0.2677 | 22.63 | 0.633336666 | -0.5445 | - | - | Variant of Uncertain Signifcance (3) | Yes | 31x | 0.93 | - |
| -0.4932 | 9.72 | 0.870912909 | -1.0613 | - | - | - | No | 31x | 0.99 | - |
| -0.1788 | 11.93 | 0.625237476 | -0.4675 | - | - | - | No | 31x | 0.81 | - |

**Supplementary Table 1:** Variant details, annotation, and interpretation. Columns report: **Patient ID**, **Chromosome**, **RefStartPos** (Reference region containing SV start position in base pairs), **RefEndPos** (Reference region containing SV end position in base pairs), **Type**, **Size** (in base pairs), **ISCN** (International System for Human Cytogenomic Nomenclature), **Overlap Genes**, **Nearest NonOverlap Gene**, **Putative Gene Fusion**, **VAF** (variant allele frequency), **VAF Supportive of Somatic Origin** (>0.35 = Ambiguous, 0.2-0.35 = Suggestive, ≤0.2 = Strongly), **Confidence**, **Frequency in Control Database**, **Number of Similar SV Type Variants in gnomAD SV** (DUP= duplications, INS= insertions, DEL=deletions, INV= inversions), **gnomAD SV Size Range** (in base pairs, kilobases, or megabases), **gnomAD SV Allele Count Range**, **Number of Enhancer-like Elements (ENCODE3 cCREs)**, **Number of Promoter-like Elements (ENCODE3 cCREs)**, **Percent Overlap SINEs (Repeat Masker)** (percentage of the reference region containing the SV overlapping SINEs) , **SINE Permutation Test p value**, **SINE Permutation Test z score**, **Percent Overlap LINEs (Repeat Masker)** (percentage of the reference region containing the SV overlapping SINEs), **LINE Permutation Test p value**, **LINE Permutation Test z score**, **Curated Lists and OMIM ID** (lists that contain the overlap or nearest non overlap genes, SysNDD, Epilepsy Gene List, and Online Mendelian Inheritance in Man), **OMIM Neurological Phenotype**, **AnnotSV ACMG** (American College of Medical Genetics) **Score of OGM Reference Regions**, **Support Found in PacBio**, **PacBio Coverage**, **Probability of ≥5 Reads in PacBio** (binomial distribution probability value), **Fit for Patient Phenotype**.

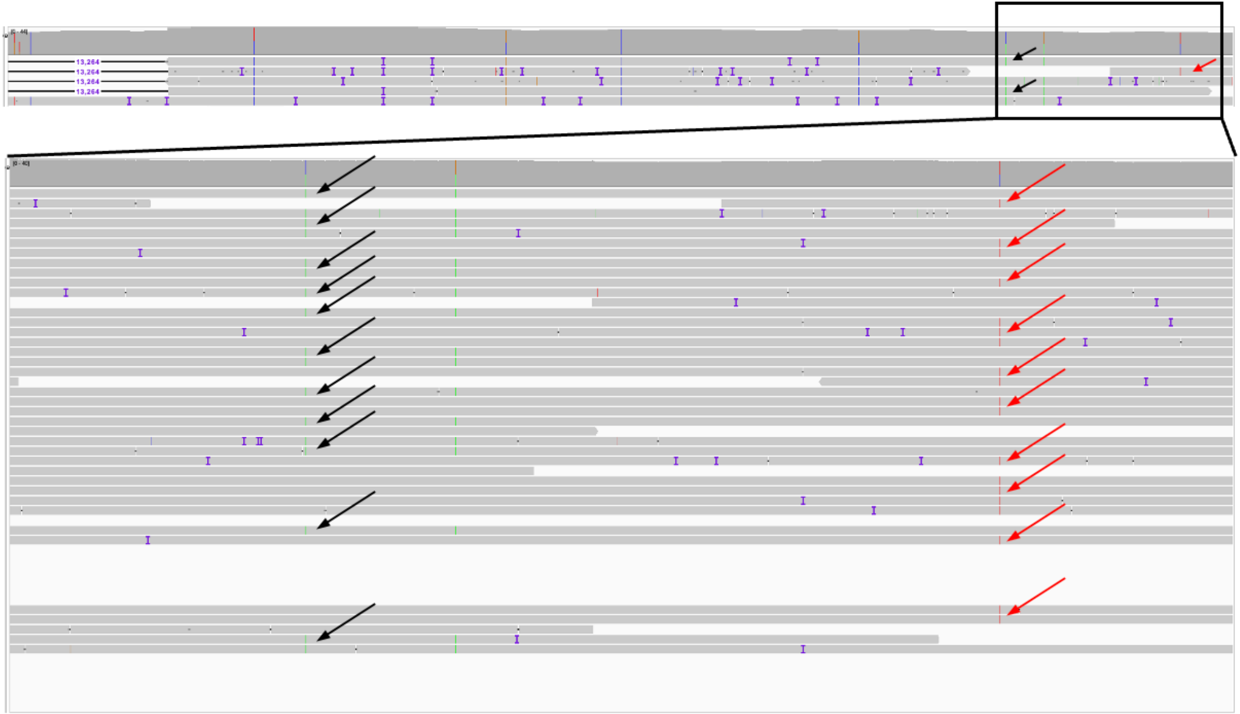

13,264
13,264
13,264
13,264

**Supplementary Figure 1:** HiFi reads spanning both the somatic deletion (far left side) and the pathogenic germline heterozygous variant in Exon 6 (far right side, red). None of the reads contained both variants, suggesting they are on opposite alleles. One SNV (green, indicated with black arrows) was found common to the somatic deletion, suggesting that it was on the same allele. Moreover, none of the reads containing this SNV were found on the same read as the heterozygous variant (red), further supporting the hypothesis that the somatic deletion occurs on the opposite allele as the germline variant.
